# Neural Networks Accurately Predict Precise Metrics of Hospital Resource Utilization for Total Hip Arthroplasty: A Retrospective Database Study

**DOI:** 10.1101/2025.02.11.25322104

**Authors:** Aazad Abbas, Johnathan R. Lex, Jay Toor, Elias B. Khalil, Bheeshma Ravi, Cari Whyne

## Abstract

**Background:** Total hip and knee arthroplasties (THAs and TKAs) are some of the most common and successful surgeries. Predicting their duration of surgery (DOS) and length of stay (LOS) has massive implications for costs and resource management. The purpose of this study was to predict the DOS and LOS of THAs using machine learning models (MLMs) based on preoperative factors.

**Methods:** The American College of Surgeons (ACS) National Surgical and Quality Improvement (NSQIP) database was queried for elective unilateral THA procedures. Multiple MLMs were constructed to predict DOS and LOS. Models were evaluated according to mean squared error (MSE), buffer accuracy, and classification accuracy. To ensure useful predictions, the results of the models were compared to a mean regressor and previous MLM predictions for primary TKAs.

**Results:** 196,942 patients were included. The neural network had the best MSE, buffer and training accuracies for both DOS and LOS. For DOS testing, the neural network MSE was 0.916, with the 30-minute buffer and ≤120 min, >120 min accuracies being 75.4% and 88.5%. For LOS testing, the neural network MSE was 0.567, with the 1-day buffer and ≤2 days, >2 accuracies being 70.3% and 80.9%. Slightly reduced performance was found for THA compared to TKA for DOS and LOS (3 to 5%), with similar important features identified.

**Conclusion:** MLMs based on preoperative factors successfully predicted the DOS and LOS of elective unilateral THAs, with similar performance to TKA. Future work should include operational factors to apply these models to real world resource optimization.

## 1 Introduction

Total hip arthroplasty arthroplasty (THA) is the fourth most performed procedure in the United States, after cesarian sections, perineal muscle laceration repair and total knee arthroplasty (TKA)[1]. Notably, THA procedures are incredibly dependable and have provided significant health and quality of life benefits to patients for decades[2–4]. Over 600,000 THA procedures are performed annually in the United States (US), with each procedure conferring significant associated costs secondary to implants, provider charges and resource utilization[5]. The demand for THA will continue to grow due to an aging population and ongoing obesity epidemic[6–9].

As surgical care already contributes to half of all hospital expenditure, this will significantly burden healthcare systems globally[10–13]. Accordingly, there are ongoing efforts to address these issues by developing clinical strategies to reduce patient length of stay (LOS) and healthcare spending[14,15]. However, despite increases in spending, demand for these procedures outweighs the supply and surgical wait times continue to increase[16]. This stresses the need for clinicians and healthcare systems to develop and leverage innovative solutions to prepare resources and ultimately aim to meet the growing demand.

Arthroplasty procedures specifically are a major contributor to these increased costs and growing waitlist[17,18]. The demand has skyrocketed for these procedures, with the ubiquity of these procedures correlating strongly with their burden on healthcare systems[17,18]. In the US, approximately 5% of the gross domestic product (GDP) is to care for musculoskeletal conditions[19,20]. As such, elective arthroplasty operations are the best candidate to focus efficiency improvement activities due to their reproducibility, prevalence, and resource-intensive nature. Despite this, little has been done to address a basic issue that drives up these costs: forecasting their demand on an individual patient basis in healthcare systems with limited resources[21,22].

A viable solution to the problem is machine learning (ML), a form of artificial intelligence (AI). In comparison to traditional statistics, ML methods are typically predictive rather than descriptive and aim at representing nonlinear relationships between features and outputs. By accurately enabling the prediction of patient-specific resource requirements, such as a patient’s duration of surgery (DOS) or length of stay (LOS), hospitals can improve the efficiency and reduce expenditure related to providing care. ML has been utilized for prediction of DOS and LOS; however, most of the current work focuses on identifying individual predictors, rather than generating comprehensive and applicable ML models[23–25]. Due to the nature of the prediction and the granularity of the datasets, these models are frequently not relevant nor generalizable.

Furthermore, these studies frequently fail to assess the accuracy of their forecasts to the current standard of care, potentially emphasizing or hiding the real effectiveness of machine learning models (MLMs). Incorporating generic patient variables into MLMs for DOS and LOS prediction can address these drawbacks and be critical towards maximizing healthcare resource allocation at the institutional and system levels, especially if built using large databases. With over 600 hospitals and millions of cases reported, the National Surgical Quality Improvement Program (NSQIP) of the American College of Surgeons (ACS) offers a premier database for solving this problem [26].

In a previous study using the NSQIP database to compare DOS and LOS predictions using preoperative factors for total knee arthroplasty (TKAs), we demonstrated improved performance of MLMs over the mean estimates for DOS and LOS[27]. The primary objective of this study was to use conventional and deep MLMs to predict the DOS and postoperative LOS of patients undergoing primary elective total hip arthroplasty based on pre-operative factors using the NSQIP database. The secondary objectives were to compare the efficacy of these models regarding the accuracy of these predictions and identify key features that may be used within these models including those generalizable for both THA and TKA.

## 2 Materials and Methods

Intuitional research ethics approval was obtained before commencement of this study (Sunnybrook Health Sciences Centre, Project ID #4899). Data is fully anonymized as per the ACS NSQIP database [26].

### 2.1 Study Population

The overall approach used is represented in Figure 1. All unilateral THA procedures completed over from 2014-2019 were sampled from ACS NSQIP, an anonymous international prospectively collated surgical database (data accessed: June 11, 2021). See Supplemental Table 1 for current procedural terminology (CPT) codes for THAs and appropriate procedures that were performed concurrently with THAs.

**Figure 1.**
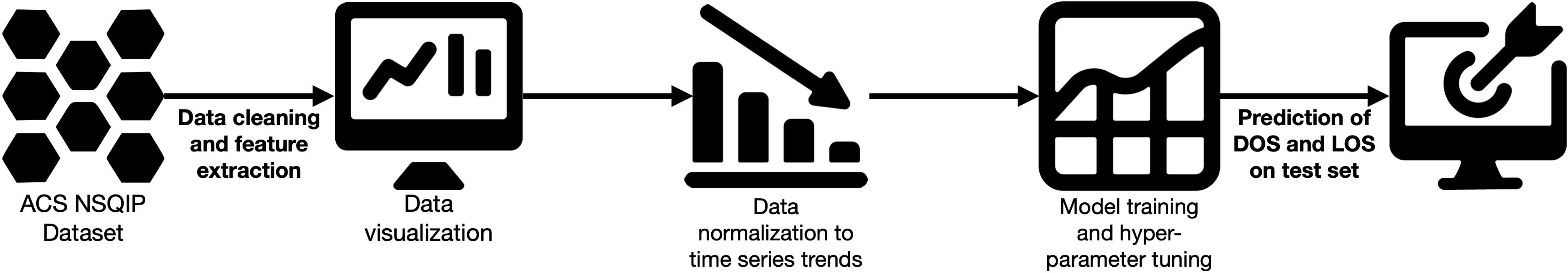
Flow diagram representing overall analysis approach utilized in this study. Data was accessed from ACS NSQIP, processed, and split, with models subsequently trained, tuned, and tested. Figure made with images from the streamline resources (https://www.streamlinehq.com/license-free) under the CC BY 4.0 license.

Excluded patients were those with an unassigned American Society of Anesthesiologists (ASA) scores, and cases with a missing or incorrectly coded DOS/LOS. Continuous variables were summarized with mean and standard deviation (SD) and categorical variables summarized with counts (N) and percentages (%). The database was split into training, validation, and testing sets by year (2014-2017 training, 2018 validation, 2019 testing).

### 2.2 Data Preprocessing

Features used to construct the models are found in Table 1. Features that were missing for more than 50% of the patients were removed, with the remaining features utilized for model inputs. To fill in missing data, a preprocessor was trained on the training set and applied to the training, validation, and test datasets. Features were appropriately scaled and adjusted. The correlation between feature pairs was determined using Pearson’s correlation coefficient.

**Table 1.** Select demographic variables of patients included in this study.

| Continuous features |  | Categorical features (binary) |  | Categorical features (multiple) |  | Ordinal features |  |
| --- | --- | --- | --- | --- | --- | --- | --- |
| Name (units) | Mean (SD) | Name | N (%) | Name | N (%) | Name | N (%) |
| Age (years) | 65.3 (11.3) | Female sex | 107893 (54.8) | Race |  | Diabetes§ |  |
| Weight (kg) | 85.9 (20.7) | Current smoker | 24548 (12.5) | White | 144037 (73.1) | No | 173011 (87.8) |
| Height (cm) | 168.1 (10.4) | On dialysis | 446 (0.2) | Unknown | 33468 (17.0) | Yes (no insulin) | 18339 (9.3) |
| Body mass index (kg/m^2) | 30.3 (6.2) | Disseminated cancer | 654 (0.3) | Black or African American | 15330 (7.8) | Yes (insulin) | 5637 (2.9) |
| Sodium (mEq/L) | 139.6 (2.9) | COPD | 7540 (3.8) | Asian | 2830 (1.4) | Dyspnea |  |
| BUN (mg/dL) | 17.8 (7.0) | CHF 30 days before procedure | 686 (0.3) | Native American/Pacific Islander | 1322 (0.7) | No | 188401 (95.6) |
| Creatinine (mg/dL) | 0.9 (0.4) | Hypertension | 108468 (55.1) | Primary Anesthesia Technique |  | Moderate exertion | 8246 (4.2) |
| Albumin (g/dL) | 4.1 (0.4) | Acute renal failure | 104 (0.05) | General | 92408 (46.9) | At rest | 340 (0.2) |
| WBCs (per µL) | 7.1 (2.3) | Steroid use* | 7315 (3.7) | Spinal | 72296 (36.7) | Functional status |  |
| Hematocrit (mg/dL) | 41.2 (4.2) | Major weight loss† | 397 (0.2) | MAC/IV Sedation | 28338 (14.4) | Independent | 193817 (98.4) |
| Platelets (per $\mu\text{L}$) | 248.6 (68.0) | Transfusion<sup>‡</sup> | 247 (0.1) | Regional | 2940 (1.5) | Partially dependent | 3040 (1.5) |
| INR | 1.03 (0.3) | Bleeding disorder | 3959 (2.0) | Epidural | 1005 (0.5) | Totally dependent | 130 (0.1) |

\*Steroid use for a chronic condition.
<sup>†</sup>Defined as >10% weight loss within 6 months of surgical procedure.
<sup>‡</sup>Defined as preoperative transfusion of $\geq 1$ unit of whole/packed RBCs in 72 hours prior to surgery.
§Diabetes status is either no diabetes, diabetes managed without insulin, or diabetes managed with insulin.
BUN: blood urea nitrogen, CHF: congestive heart failure, COPD: chronic obstructive pulmonary disease, INR: international normalized ratio, WBCs: white blood cells.

DOS and LOS for each dataset was normalized to adjust for the change by year, with a regression model based on the training data used to adjust the validation and test data. A log-normal distribution transformation was applied to DOS and LOS to allow for optimal MLM results[28,29]. Additional details on these methods are provided in Abbas et al 2022[27].

### 2.3 MLM Construction

A multi-layer perceptron (MLP) was used as the deep learning model[30]. An MLP is an artificial neural network (ANN) that contains an input layer, hidden layers, and an output layer. The features are described in the input layer, the hidden layer(s) are the nonlinear interactions between features, and the output layer is models’ prediction. Conventional MLMs used were linear regression, stochastic gradient descent (SGD) linear regression, K-nearest neighbors (KNN), decision trees, random forest, AdaBoost, elastic net, and linear support vector machine (SVM) regression. A mean regressor was built to compare the MLMs to a non-predictive model, i.e., a model that uses the mean of the training data as the result for the test data.

Models were trained and hyperparameters tuned to minimize the mean squared error (MSE), with the analysis done on the Mist supercomputer at the SciNet HPC Consortium using the Tune package (Supplemental Table 2) [31–33]. When possible, feature importance was extracted from the models and normalized with respect to the most important feature for each model in percentages.

### 2.4 Outcome Metrics

MSE, buffer accuracy, and classification accuracy were used to evaluate each model as interpretable outcome metrics for the predictions, with the quality of the final models evaluated on the test set[27]. Buffer accuracy is defined as how often the predicted DOS/LOS was within a predesignated value of the actual DOS/LOS, while classification accuracy is defined as the percentage of times the correct classification group value was predicted by the models. Buffer accuracy for DOS was measured at 15-minute intervals ranging from 15 minutes to 60 minutes, while classification accuracy for DOS was measured in three groups: 1) ≤ 90 minutes, > 90 minutes, 2) ≤120 minutes, >120 minutes, and 3) <60 minutes, 60 to 90 minutes, > 90 minutes.

Buffer accuracy for LOS was measured at 1 day, 1.5 days, and 2 days while classification accuracy was measured in three groups: 1) ≤ 1 day, >1 day, 2) ≤2 days, >2 days, and 3) <1 day, 1 to 3 days, > 3 days. A description of these metrics is provided in Abbas et al[27]. Statistical analysis was conducted using Python (Python Software Foundation, www.python.org)[34,35].

## 3 Results

### 3.1 Patient Demographics

A total of 196,987 patients were included, with 117,460 in the training set, 37,955 in the validation set, and 41,572 in the testing set. Thirty-three features were used to construct the models. The mean DOS and LOS were 90.2 (standard deviation (SD) 36.8) minutes and 2.3 (SD 2.4) days. Figure 2 displays a network plot of the correlation between all the features used to train the models. Table 1 provides a breakdown of patient demographics.

**Figure 2.**
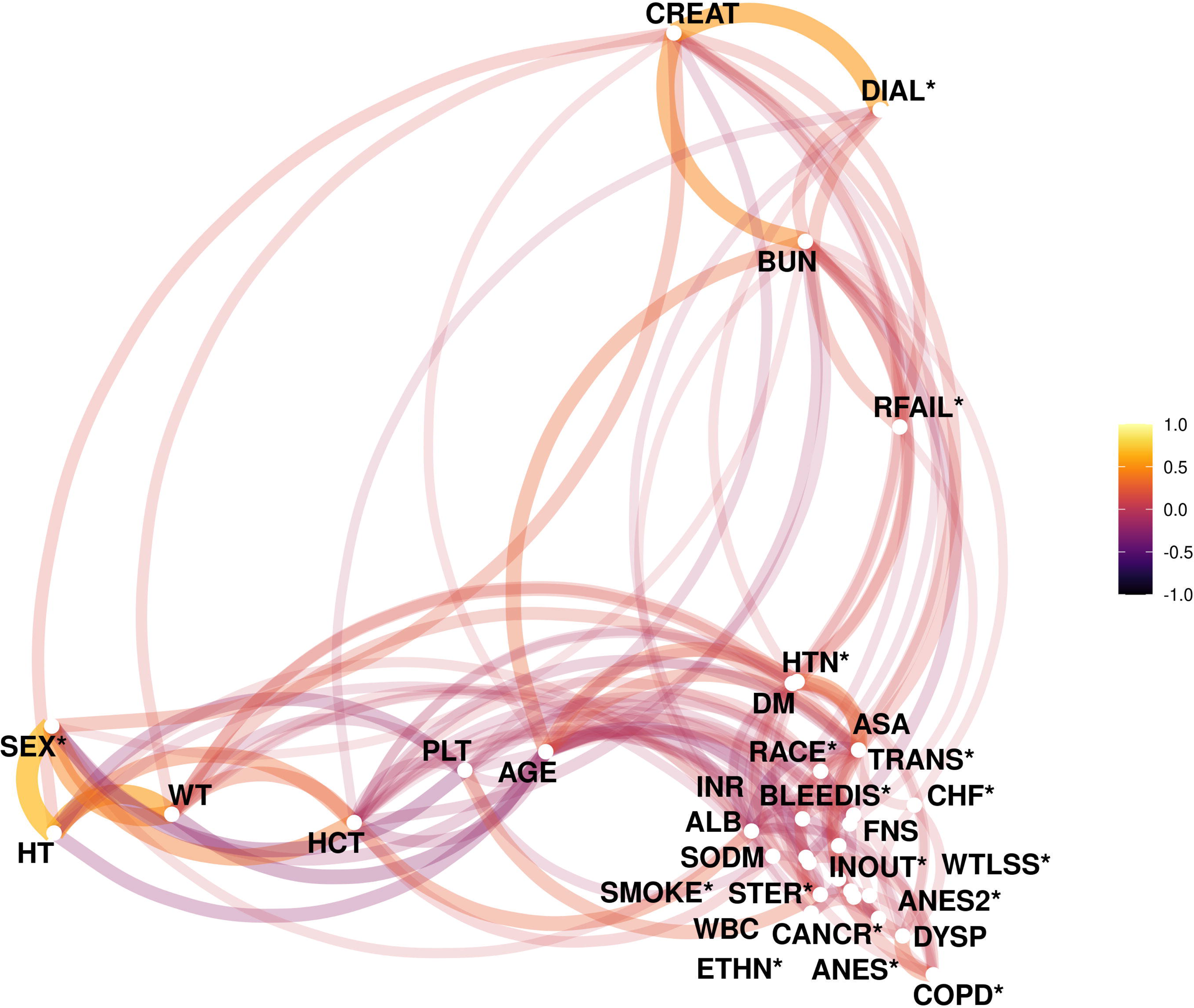
Network plot representing the correlation between features in the training set. Pearson correlations coefficients with an absolute value of greater than 0.05 are shown. Line thickness and proximity represents strength of correlation. Colour represents sign of correlation. *Categorical features. ALB: albumin (g/dL), ANES: primary anesthesia technique, ANES2: secondary anesthesia technique, ASA: American Society of Anesthesiologists classification, BLEEDIS: bleeding disorder, BUN: blood urea nitrogen (mg/ dL), CANCR: disseminated cancer, CHF: congestive heart failure status, COPD: chronic obstructive pulmonary disease status, CREAT: creatinine (mg/dL), DIAL: currently on dialysis, DM: diabetes status, DYSP: dyspnea status, ETHN: ethnicity (Hispanic), FNS: functional status, HCT: hematocrit (mg/dL), HT: height, HTN: hypertension status, INOUT: inpatient/outpatient status, INR: international normalized ratio, PLT: platelets (per μL), RFAIL: renal failure status, SMOKE: smoking status, SODM: sodium (mEq/L), STER: steroid use, TRANS: preoperative transfusion within 72 h, WBC: white blood cells count (per μL), WT: weight (kg), WTLSS: >10% weight loss in last 6 months.

### 3.2 Model Results

Results of training the DOS and LOS models are shown in Tables 2 and 3, while results of validation and testing of the DOS and LOS models are shown in Tables 4 and 5.

**Table 2.** Results of the training set for the duration of surgery (DOS) predictions. Accuracies represented in percentages.

| Model Name | MSE | Buffer<br>accuracy<br>(15 min) | Buffer<br>accuracy<br>(30 min) | Buffer<br>accuracy<br>(45 min) | Buffer<br>accuracy<br>(60 min) | Classification<br>accuracy<br>group 1* | Classification<br>accuracy<br>group 2† | Classification<br>accuracy<br>group 3‡ |
| --- | --- | --- | --- | --- | --- | --- | --- | --- |
| Mean Regressor | 1 | 35.6 | 66.4 | 85.6 | 93.7 | 42.7 | 83.5 | 42.7 |
| Linear Regression | 0.951 | 41.2 | 71.6 | 87.2 | 93.2 | 61.9 | 83.5 | 46.9 |
| SGD Regression | 0.948 | 40.6 | 71.0 | 87.2 | 93.4 | 61.7 | 83.5 | 47.4 |
| KNN | 0.772 | 46.8 | 76.5 | 89.5 | 94.7 | 69.3 | 84.1 | 54.5 |
| Decision Tree | 0.011 | 97.1 | 98.3 | 99.9 | 100 | 98.1 | 99.2 | 99.9 |
| Random Forest | 0.219 | 81.6 | 94.3 | 97.7 | 98.9 | 92.9 | 90.7 | 83.7 |
| AdaBoost | 0.974 | 40.7 | 70.9 | 86.9 | 93.0 | 60.3 | 83.5 | 44.7 |
| Elastic Net | 1.037 | 39.0 | 69.0 | 86.2 | 92.4 | 57.4 | 83.5 | 40.2 |
| Linear SVM | 0.958 | 40.8 | 71.2 | 87.0 | 93.2 | 61.6 | 83.5 | 46.9 |
| Neural Network | 0.888 | 43.7 | 74.0 | 89.0 | 94.8 | 65.5 | 84.8 | 50.6 |

\*Classification accuracy group 1 represents classification of $\leq 90$ minutes, $> 90$ minutes.
†Classification accuracy group 2 represents classification of $\leq 120$ minutes, $> 120$ minutes.
‡Classification accuracy group 3 represents classification of $\leq 60$ minutes, 60-90 minutes, $> 90$ minutes.
KNN: K-nearest neighbour, MSE: mean squared error, SGD: stochastic gradient descent, SVM: support vector machine.

**Table 3.** Results of the training sets for the length of stay (LOS) predictions. Accuracies represented in percentages.

| Model Name | MSE | Buffer accuracy<br>(1 day) | Buffer accuracy<br>(1.5 days) | Buffer accuracy<br>(2 days) | Classification<br>accuracy group<br>1* | Classification<br>accuracy group<br>2† | Classification<br>accuracy group<br>3‡ |
| --- | --- | --- | --- | --- | --- | --- | --- |
| Mean Regressor | 1 | 62.0 | 83.4 | 91 | 91.9 | 60.7 | 65.3 |
| Linear Regression | 0.972 | 70.3 | 86.1 | 91.8 | 92 | 66.3 | 67.8 |
| SGD Regression | 1.254 | 70.1 | 86 | 91.7 | 92 | 66.5 | 67.7 |
| KNN | 0.842 | 75.2 | 88.5 | 93.1 | 92 | 71.9 | 70.3 |
| Decision Tree | 0.012 | 97.3 | 98.1 | 99.8 | 97.8 | 98.2 | 99.4 |
| Random Forest | 0.465 | 93 | 95.8 | 97.1 | 95.3 | 92.9 | 83.1 |
| AdaBoost | 0.969 | 68.7 | 85.9 | 91.2 | 91.9 | 65.5 | 66.6 |
| Elastic Net | 1.028 | 65.6 | 86.1 | 90.6 | 91.9 | 60.7 | 65.3 |
| Linear SVM | 0.998 | 70.2 | 86 | 91.9 | 92 | 67 | 67.8 |
| Neural network | 0.860 | 72.6 | 87.4 | 92.6 | 92.1 | 69 | 69.3 |

\*Classification accuracy group 1 represents classification of $\leq 1$ day, 1 day +.
†Classification accuracy group 2 represents classification of $\leq 2$ days, 2 days +.
‡Classification accuracy group 3 represents classification of $< 1$ day, 1-3 days, 3 days +.
KNN: K-nearest neighbor, MSE: mean squared error, SGD: stochastic gradient descent, SVM: support vector machine.

**Table 4.** Results of the validation and testing sets for the duration of surgery (DOS) predictions. Accuracies represented in percentages.

| Model Name | MSE | Buffer<br>accuracy<br>(15 min) | Buffer<br>accuracy<br>(30 min) | Buffer<br>accuracy<br>(45 min) | Buffer<br>accuracy<br>(60 min) | Classification<br>accuracy<br>group 1* | Classification<br>accuracy<br>group 2† | Classification<br>accuracy<br>group 3‡ |
| --- | --- | --- | --- | --- | --- | --- | --- | --- |
| Validation Set |  |  |  |  |  |  |  |  |
| Mean Regressor | 1.08 | 37.4 | 68.8 | 87.6 | 94.2 | 59.4 | 85.4 | 41.3 |
| Linear Regression | 1.048 | 42.1 | 73.5 | 88.1 | 93.5 | 61.9 | 85.4 | 45.3 |
| SGD Regression | 1.052 | 41.7 | 72.9 | 88.1 | 93.6 | 61.8 | 85.3 | 45.7 |
| KNN | 1.167 | 39.3 | 69.1 | 85.1 | 92.3 | 58.9 | 84.5 | 43.8 |
| Decision Tree | 1.984 | 30.9 | 56.6 | 73.7 | 84.6 | 55.4 | 76.2 | 39.7 |
| Random Forest | 1.057 | 41.8 | 72.7 | 87.8 | 93.4 | 61.4 | 85.4 | 45.3 |
| AdaBoost | 1.079 | 41.8 | 72.8 | 87.7 | 93.2 | 60.4 | 85.4 | 43 |
| Elastic Net | 1.131 | 40.1 | 71.6 | 87.2 | 92.8 | 59.4 | 85.4 | 41.3 |
| Linear SVM | 1.056 | 41.7 | 72.9 | 88 | 93.5 | 61.8 | 85.4 | 45.4 |
| Neural network | 0.910 | 45.2 | 75.0 | 89.4 | 95.5 | 63.2 | 87.2 | 47.3 |
| Testing Set |  |  |  |  |  |  |  |  |
| Mean Regressor | 1.182 | 37.5 | 68.6 | 87.5 | 94.1 | 59.5 | 85.6 | 40.7 |
| Linear Regression | 1.16 | 42.9 | 73.2 | 87.7 | 93.2 | 62.3 | 85.6 | 44.6 |
| SGD Regression | 1.147 | 42.3 | 73 | 87.8 | 93.4 | 62.3 | 85.6 | 44.5 |
| KNN | 1.272 | 40 | 69.3 | 85.2 | 92.2 | 59.8 | 85.1 | 42.8 |
| Decision Tree | 2.047 | 31.7 | 57.5 | 74.8 | 85.4 | 56.4 | 78.2 | 39.3 |
| Random Forest | 1.166 | 42.6 | 72.9 | 87.5 | 93.1 | 61.8 | 85.6 | 43.9 |
| AdaBoost | 1.182 | 41.9 | 72.8 | 87.6 | 93.1 | 60.3 | 85.6 | 41.6 |
| Elastic Net | 1.242 | 40.2 | 71.2 | 87 | 92.7 | 59.5 | 85.6 | 40.7 |
| Linear SVM | 1.161 | 42.5 | 73 | 87.6 | 93.2 | 62.3 | 85.6 | 44.1 |
| Neural network | 0.916 | 45.1 | 75.4 | 90.2 | 96 | 65.2 | 88.5 | 47.5 |
| Neural network – TKA<br>[27] | 0.893 | 46.5 | 78.4 | 91.4 | 96.1 | 62.4 | 87.7 | 48.4 |

\*Classification accuracy group 1 represents classification of $\leq 90$ minutes, $> 90$ minutes.
†Classification accuracy group 2 represents classification of $\leq 120$ minutes, $> 120$ minutes.
‡Classification accuracy group 3 represents classification of $\leq 60$ minutes, 60-90 minutes, $> 90$ minutes.
KNN: K-nearest neighbor, MSE: mean squared error, SGD: stochastic gradient descent, SVM: support vector machine.

**Table 5.**
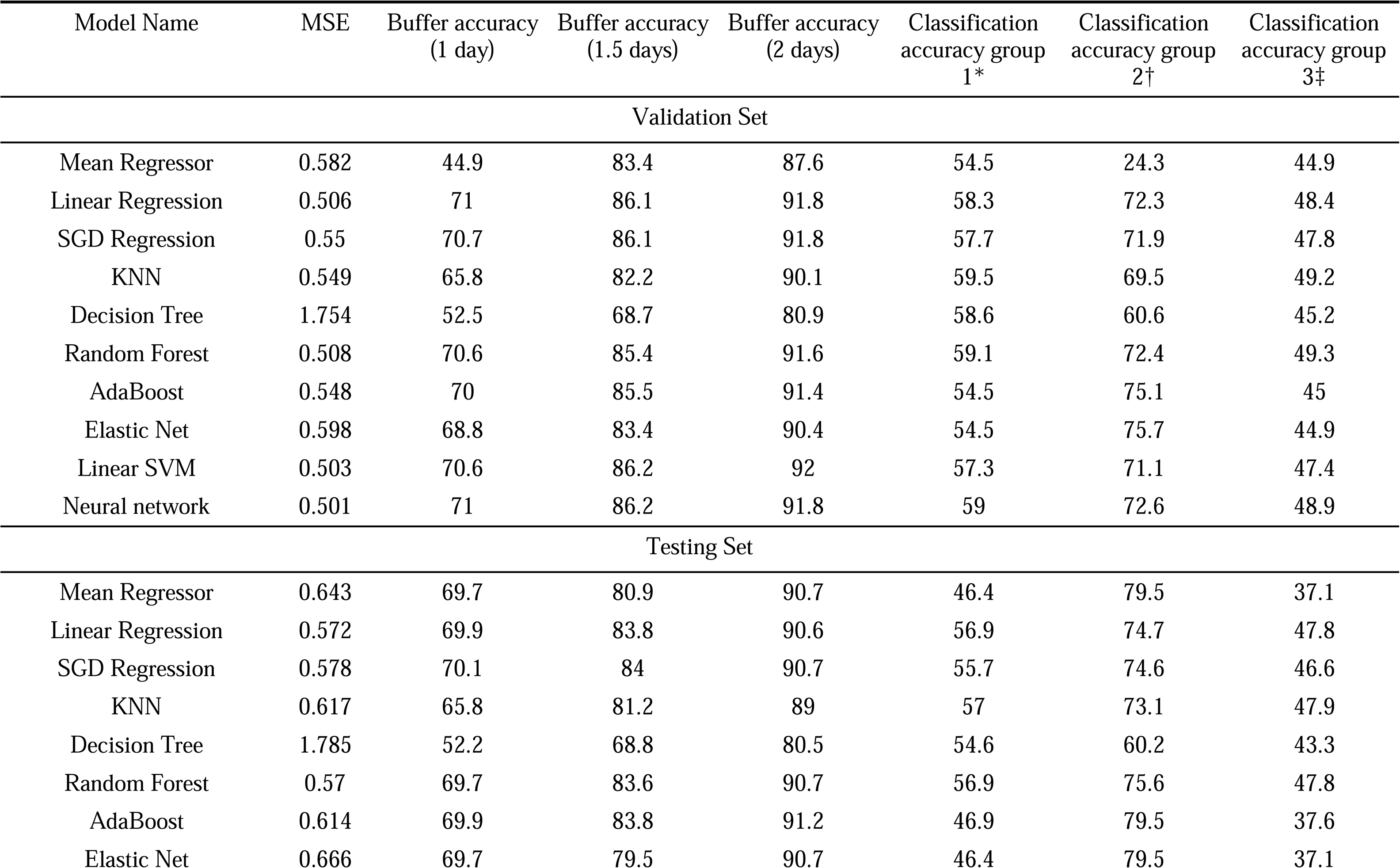
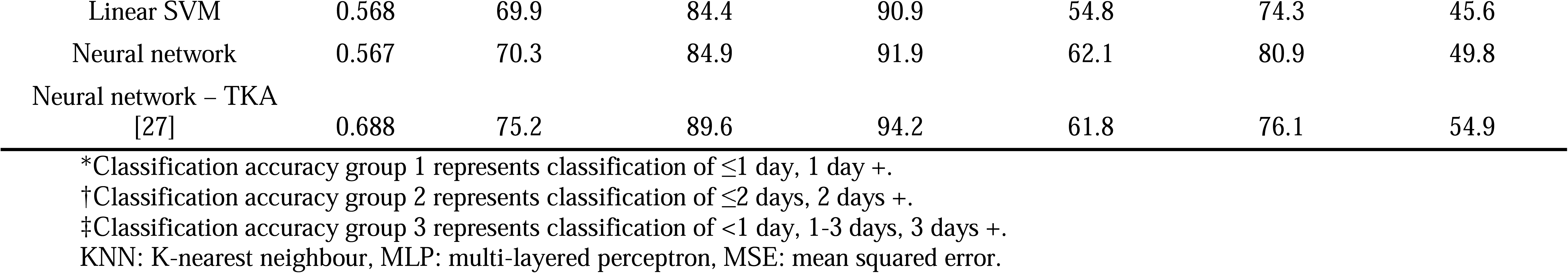
Results of the validation and testing sets for the length of stay (LOS) predictions. Accuracies represented in percentages.

#### 3.1.1 Model Training

For both DOS and LOS, the decision tree model resulted in the lowest training MSE of 0.011 and 0.012, respectively. Similarly, the decision tree model had near 100% accuracy for both DOS and LOS in training (Tables 2 and 3). For both DOS and LOS, the random forest model was second to decision trees by MSE, buffer and classification accuracies (Tables 2 and 3). For DOS and LOS, the KNN model had the third lowest MSE and third highest buffer and classification accuracies (Table 2 and 3). All models outperformed the mean regressor for both buffer and classification accuracies during training.

#### 3.1.2 Model Validation and Testing

For DOS validation, the neural network produced the lowest MSE of 0.910, as well as the highest 15- and 30-minute buffer accuracies of 45.2% and 75.0%, respectively. In addition, all the models outperformed the mean regressor by 2-8% across all outcome metrics. The KNN, decision, tree, and elastic net models MSEs were higher than the mean regressor, i.e., worse models, which was reflected in their corresponding accuracy metrics. Similarly for DOS testing, the neural network was the superior model via an MSE of 0.916 and higher buffer and classification accuracies (Table 4). The 15- and 30- minute buffer accuracies were 45.1% and 75.4% with the classification accuracy group 2 being 88.5% (Table 4). Again, the KNN, decision tree, and elastic net models performed worse than the mean regressor via MSE metrics, with the decision tree model consistently performing worse than the mean regressor according to accuracy metrics.

For LOS validation, the neural network produced the lowest MSE of 0.501 with the linear regression model resulting in similar 1- and 2-day buffer accuracies of 71% and 91.8% (Table 5). Compared to the mean regressor, the accuracies of the models were 2% to 48% better. The decision tree and elastic net models MSEs were worse than the mean (Table 5). Similarly for LOS testing, the neural network was the best performing model with an MSE of 0.567 and higher buffer and classification accuracies (Table 5). Namely, the 1- and 2-day buffer accuracies were 70.3% and 91.9% with the classification accuracy group 2 being 80.9% (Table 5).

### 3.2 Feature Importance

The top ten features for select models for DOS and LOS are displayed in Figures 3 and 4, respectively.

**Figure 3.**
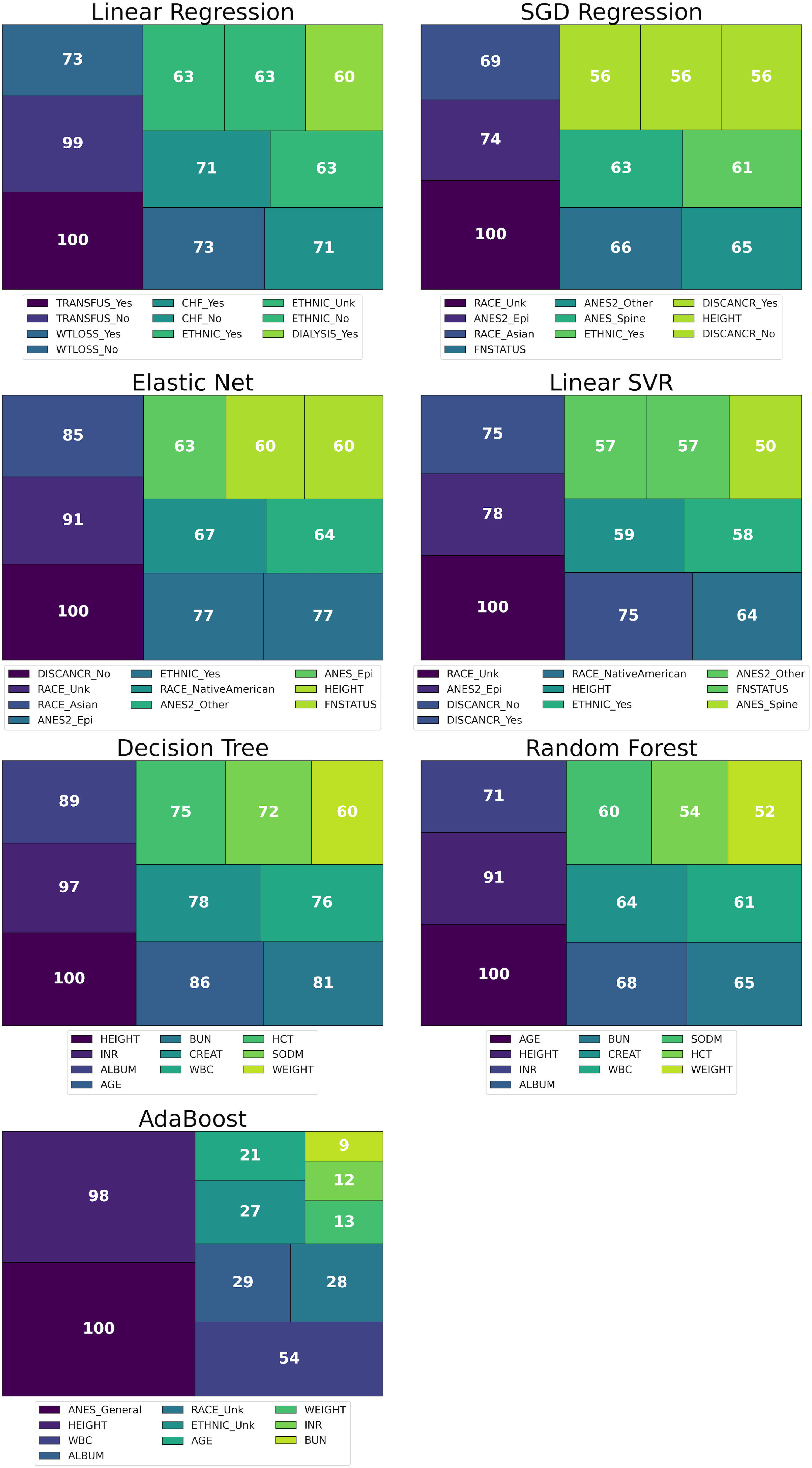
Tree plots representing the top ten most important features for linear and tree-based models for the duration of surgery prediction. Numbers indicate normalized feature importance in percentages, with legends below each plot corresponding to respective features. ALBUM: albumin (g/dL), ANES: primary anesthesia technique, ANES2: secondary anesthesia technique, BUN: blood urea nitrogen (mg/ dL), CHF: congestive heart failure status, CREAT: creatinine (mg/dL), DISCANCR: disseminated cancer, ETHNIC: ethnicity (Hispanic), FNSTATUS: functional status, HCT: hematocrit (mg/dL), INOUT: inpatient/outpatient status, INR: international normalized ratio, PLATE: platelets (per μL), SODM: sodium (mEq/L), TRANSFUS: preoperative transfusion within 72 h, WBC: white blood cells count (per μL), WTLSS: >10% weight loss in last 6 months.

**Figure 4.**
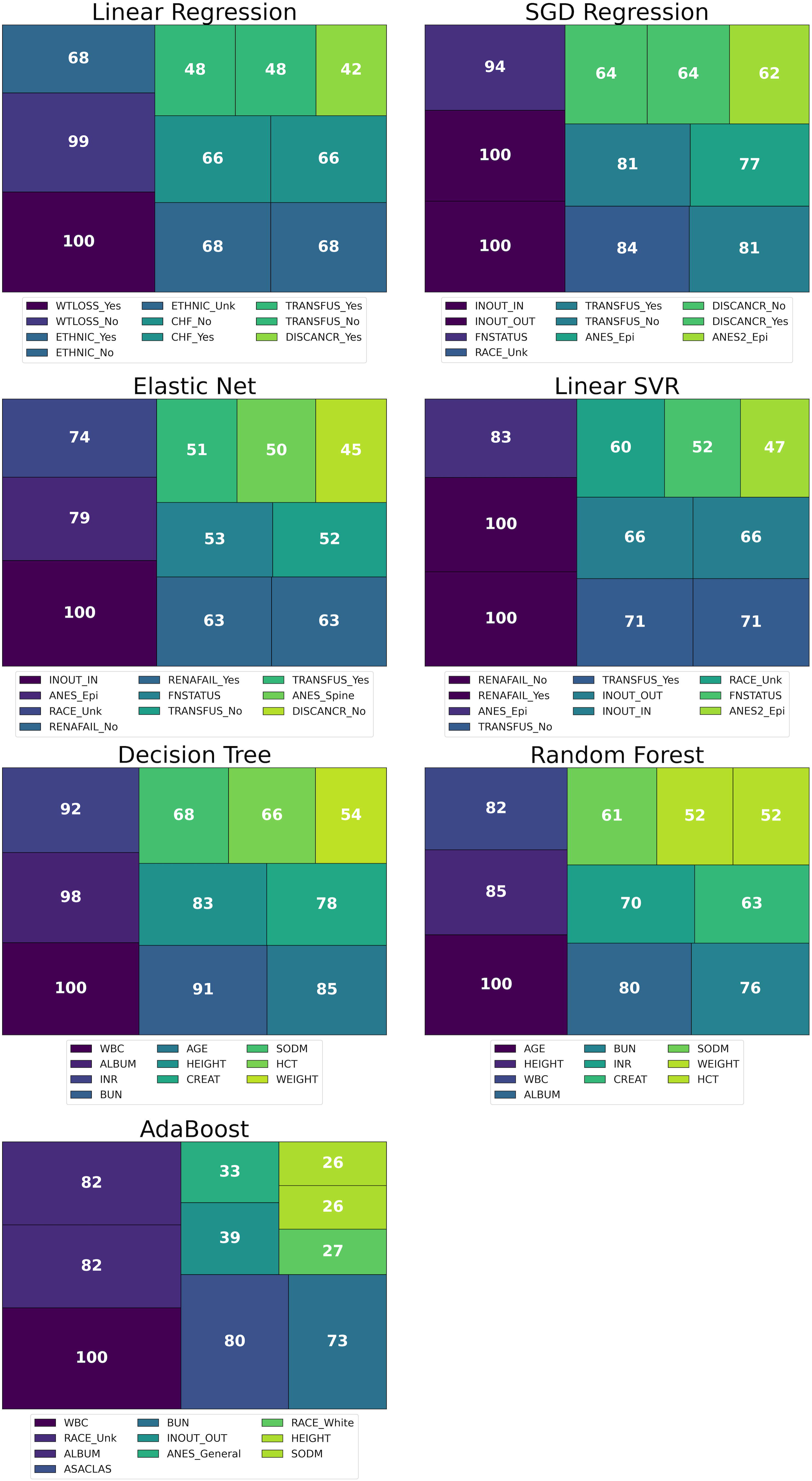
Tree plots representing the top ten most important features for linear and tree-based models for the length of stay predictions. Numbers indicate normalized feature importance in percentages, with legends below each plot corresponding to respective features. ALBUM: albumin (g/dL), ANES: primary anesthesia technique, ANES2: secondary anesthesia technique, ASA: American Society of Anesthesiologists classification, BUN: blood urea nitrogen (mg/ dL), CHF: congestive heart failure status, COPD: chronic obstructive pulmonary disease status, CREAT: creatinine (mg/dL), DIAL: currently on dialysis, DISCANCR: disseminated cancer, ETHNIC: ethnicity (Hispanic), FNSTATUS: functional status, HCT: hematocrit (mg/dL), INOUT: inpatient/outpatient status, INR: international normalized ratio, PLATE: platelets (per μL), RENAFAIL: renal failure status, SODM: sodium (mEq/L), TRANSFUS: preoperative transfusion within 72 h, WBC: white blood cells count (per μL), WTLSS: >10% weight loss in last 6 months.

Linear models for both DOS and LOS placed greater importance on features such as pre-existing conditions (e.g., renal failure, CHF status, disseminated cancer), patient demographic characteristics like race and sex, and preoperative factors including type of anesthesia used and inpatient/outpatient status. While including some of the features shared with the linear models, the tree-based models placed a larger emphasis on numerical features namely preoperative lab values, weight, height, and age.

The top features for the best conventional MLM for DOS and LOS prediction — linear regression — were transfusion given within 72 hours preoperatively, weight loss in last 6 months, CHF status, ethnicity, and currently receiving dialysis.

### 3.3 Performance and Feature Comparison to TKA Models

Performance of conventional and deep MLMs in predicting DOS and LOS was comparable for THA to that previously found for TKA (Tables 4, 5) [27]. In particular, the best DOS 30-minute buffer accuracy (THA 75.4% vs TKAs 78.8%) and LOS 1-day buffer accuracy (THA 70.3% vs TKAs 75.2%) were similar, with performance superior for TKA by 3-5% for both outcomes. In considering the fit of the models, THA neural networks performed worse than TKA for DOS (MSE 0.916 vs 0.896) but better for LOS (MSE 0.567 vs 0.690).

Features considered important to the models were consistent between both THA and TKA. Linear models considered pre-existing conditions, patient demographics and preoperative factors the most important while tree-based models emphasized factors such as preoperative lab values, patient weight, height, and age.

## 4 Discussion

This study used MLMs to successfully predict the DOS and postoperative LOS for primary elective unilateral THA, with neural networks shown to be the best performing model. The degree of detail generated in this paper offers significant insights into the viability of MLMs to predict outcomes that dictate healthcare resource usage.

As expected, the neural network was the best overall model for both DOS and LOS predictions. This was echoed in the various accuracy metrics of the models, with the DOS 30-minute buffer and ≤120 minutes, >120 minutes classification accuracies being 75.4% and 88.5% and the LOS 2-day buffer and the ≤2 days, 2 days + classification accuracies of 91.8% and 80.9%, respectively. The neural network developed is a predictive model composed from multiple layers of fully-connected variables. However, more traditional ML models such as the KNN, decision tree and elastic net performed worse than the mean regressor for DOS predictions, with the decision tree and elastic net models also performing worse for the LOS predictions. This is of interest, as it is often the misconception that MLMs are superior to traditional predictive statistical methods and would consistently outperform a mean value estimate. Construction of MLMs is resource intensive, so this result highlights the importance of knowing which MLMs may be most useful for a particular problem.

The implications of this work are significant. Firstly, being able to accurately predict the DOS and LOS with this level of granularity allows one to schedule surgeries more accurately. This work goes beyond previous studies whereby postoperative LOS was predicted using arbitrary cut-offs, as predicting DOS and LOS as continuous targets allows for scheduling surgery to the minute and assigned bed capacity to the day[24,25,36–38]. Secondly, this gives one the capacity to appropriately allocate staffing requirements needed in the operating room and the ward.

Applying models which better predict DOS and LOS to local hospital scheduling has the potential for massive savings by reducing staff overtime hours and facilitating appropriate resource allocation. Thirdly, by efficiently scheduling the surgeries according to what neural networks predict, hospitals can reduce overtime and/or increase throughput and thereby potentially reduce surgical waitlists. Improvements in scheduling and optimization have the potential to reduce healthcare costs, on a local institutional and health systems level. Due to the frequency and cost of arthroplasty procedures, minor improvements in the efficiency of care can have large realizable benefits for hospitals and patients.

In an era of increasing patient demand for THA and pressure for hospitals and healthcare systems to reduce the cost of delivered care, leveraging novel strategies and technology such as neural networks will be important[1,6,10,39]. Neural network models trained on large datasets have the advantages of being generalizable as they have been exposed to many examples.

Moreover, neural networks also maintain the ability to generate predictions very specific at an individual level due to their high level of complexity. The features models identified as most important for predicting both DOS and LOS were those such as patient demographics and laboratory tests that are routinely collected preoperatively (e.g., electrolytes, prothrombin time, albumin etc.). This is important to help inform variables that should be included when generating institution-specific models or implementing these models.

The construction of these models is resource intensive and mathematically challenging. This and the previous study related to TKAs was conducted using a large, internationally renowned surgical database all the while utilizing a supercomputer with cutting edge machine learning software[26,27,34,35]. Despite the advantages of this dataset, utilizing data generated from hundreds of institutions of varying sizes and clinical practices increases the complexity and heterogeneity of the dataset and its analysis. Primary joint replacements such as THAs and TKAs are very homogenous surgeries with respect to approach, surgical technique, and operating room practices. This helps make these procedures more efficient but makes it more challenging to accurately predict their DOS or LOS based on patient features. This may highlight the importance of including certain operational factors available from institutional databases and not necessarily captured in national databases such as NSQIP. Other procedures with larger variance of DOS and LOS, such as revision arthroplasty or trauma cases, may be more amenable to such predictions based on these patient-related features.

Compared to the previous study which focused on using conventional and deep MLMs for TKAs, this study showed similar outcome results with slightly reduced performance for THAs. This may be explained by a few factors; TKAs have less variability in factors that aren’t captured in the NSQIP database such as surgical approach, patient positioning, and implant types, which may allow for better fitting models for DOS. Additionally, hospital-specific factors such as surgical team structure and experience may have a larger effect on the DOS of THAs, making their outcomes harder to predict. As the MSE of the neural network MLM for LOS was lower for THA, and the NSQIP data primarily pertained to patient and anaesthetic factors, this may suggest these are more strongly correlated with LOS following THA compared to TKA.

Despite the detailed analysis, this study has some limitations. Firstly, only patient factors were used in this study. This may greatly limit maximum accuracy of the DOS and LOS predictions. Incorporating institutional factors, such as hospital infrastructure, surgical team structure and surgeon training, into the modeling may further improve DOS and LOS predictions. Secondly, despite the ACS NSQIP database being large and detailed, there is variability in the quality and accuracy of data recording[40]. As such, this may affect the quality of the models generated and subsequently the accuracy of their predictions. For example, the performance of the tree-based models was likely due to overfitting of the models. This occurred despite hyperparameter tuning and branch and depth limitations set during hyperparameter tuning on the validation set.

In conclusion, this study has utilized conventional and deep MLMs to predict DOS and LOS for unilateral THAs using preoperative factors. Multiple statistical and forecasting practices were compared, and these outcomes were predicted most accurately using neural. Notably, with these factors, LOS prediction is superior to DOS prediction, highlighting the importance of feature and dataset selection when aiming to predict specific outcomes. Studies in the future should aim to incorporate institutional factors and test the real-world efficacy of these models at improving care efficiency through prospective clinical trials.

## Data Availability

Data is fully anonymized as per the American College of Surgeons NSQIP database. Data is available upon request from them at.

## Acknowledgements

Database: The American College of Surgeons National Surgical Quality Improvement Program and the hospitals participating in the ACS NSQIP are the source of the data used herein; they have not verified and are not responsible for the statistical validity of the data analysis or the conclusions derived by the authors.

Computations were performed on the Mist supercomputer at the SciNet HPC Consortium. SciNet is funded by: the Canada Foundation for Innovation; the Government of Ontario; Ontario Research Fund - Research Excellence; and the University of Toronto.

