## Supplemental Table 1 for "Neural Networks Accurately Predict Precise Metrics of Hospital Resource Utilization for Total Hip Arthroplasty: A Retrospective Database Study"

Supplemental Table 1. CPT Codes for procedures listed concurrently with total hip arthroplasty.

| CPT Code | Procedure name |
| --- | --- |
| 27000 | Tenotomy, adductor of hip, percutaneous (separate procedure) |
| 27001 | Tenotomy, adductor of hip, open |
| 27005 | Tenotomy, hip flexor(s), open (separate procedure) |
| 27006 | Tenotomy, abductors and/or extensor(s) of hip, open (separate procedure) |
| 27033 | Arthrotomy, hip, including exploration or removal of loose or foreign body |
| 27054 | Arthrotomy with synovectomy, hip joint |
| 27086 | Removal of foreign body, pelvis or hip; subcutaneous tissue |
| 27087 | Removal of foreign body, pelvis or hip; deep (subfascial or intramuscular) |
| 27390 | Tenotomy, open, hamstring, knee to hip; single tendon |
| 27391 | Tenotomy, open, hamstring, knee to hip; multiple tendons, 1 leg |
