## Supplemental Table 2 for "Neural Networks Accurately Predict Precise Metrics of Hospital Resource Utilization for Total Hip Arthroplasty: A Retrospective Database Study"

Supplemental Table 2. Hyperparameters of duration of surgery and length of stay models.

| Model Name | Hyperparameters | |
| --- | --- | --- |
| Duration of Surgery | | |
| Mean Regressor | | No hyperparameters |
| Linear Regression | | fit_intercept=True, normalize =True |
| SGD Regression | | loss='epsilon_insensitive', penalty='l1', fit_intercept=True, alpha=0.0014595194618015974, l1_ratio=0.12371916051592253, tol=0.0005885960824465063, epsilon=0.2828473287077091, eta0=3.6413395752926386e-05, power_t=0.17345091203452156, validation_fraction=0.1, n_iter_no_change=10 |
| Elastic Net | | alpha=1.9682317264449496e-05, l1_ratio=0.9560803380368482, random_state=1, tol=2.1793498818776042e-05, fit_intercept=False, normalize=True, selection='cyclic', warm_start=True |
| Linear SVM | | tol=1e-4, C=1.0, loss='epsilon_insensitive', fit_intercept=True, intercept_scaling=1, max_iter=1000 |
| KNN | | n_neighbors=5, weights='uniform', algorithm='auto', leaf_size=30 |
| Decision Tree | | min_samples_split=2, min_samples_leaf=2, max_features='sqrt', max_depth=6 |
| Random Forest | | n_estimators=200, min_samples_split=2, min_samples_leaf=1, bootstrap=True, oob_score=False, |
| AdaBoost | | n_estimators=200, learning_rate=1.0 |
| XGBoost | | objective='reg:squarederror', n_estimators=50, eta=0.3, max_depth=6, reg_lambda=1 |
| Neural network | | learning_rate_init=7.018000343964287e-05, hidden_layer_sizes=(100, 200), alpha=6.415884013271812e-06, beta_1=0.7858927938713562, learning_rate='adaptive', beta_2=0.8916607095443794, batch_size=256,activation='relu', solver='adam' |
| Length of Stay | | |
| Mean Regressor | | No Hyperparameters |
| Linear Regression | | fit_intercept=False, normalize =False |
| SGD Regression | | loss='epsilon_insensitive', penalty='elasticnet', alpha=0.0009458700020840921, l1_ratio=0.3833695731927218, fit_intercept=True, tol=0.00015213841254039473, epsilon=0.3424422770265162, learning_rate='optimal', eta0=0.0009697535786895821, power_t=0.153079193374453 |
| Elastic Net | | alpha=1.1349632230271373e-05, l1_ratio=0.7461854674829281, fit_intercept=False, normalize=True, tol=7.354389414625075e-07, random_state=1, warm_start=True |
| Linear SVM | | tol=1e-4, C=1.0, loss='epsilon_insensitive', fit_intercept=True, intercept_scaling=1 |
| KNN | | n_neighbors=5, weights='uniform', algorithm='auto', leaf_size=30 |
| Decision Tree | | min_samples_split=2, min_samples_leaf=2, min_weight_fraction_leaf=0.0, max_features='sqrt', max_depth=6 |
| Random Forest | | n_estimators=100, min_samples_split=2, min_samples_leaf=1, max_features='auto', bootstrap=True, oob_score=False |
| AdaBoost | | n_estimators=100, learning_rate=1.0, loss="linear" |
| XGBoost | | objective='reg:squarederror', n_estimators=50, eta=0.3, max_depth=6, reg_lambda=1 |
| PyTorch MLP | | activation='relu', alpha=5.921762581814381e-06, batch_size=32, beta_1=0.8420849590775347, beta_2=0.7626345457352179, learning_rate='adaptive', hidden_layer_sizes=(100, 100), learning_rate_init=0.0004985820888366887 n_iter_no_change=10, solver='adam' |
